# Fight or Flight: Emergency Healthcare Workers’ Willingness to Work during Crises and Disasters: A cross-sectional multicentre study in the Netherlands

**DOI:** 10.1101/2023.07.25.23293139

**Authors:** Lindsy M.J. Engels, Dennis G. Barten, Thimo J.T. Boumans, Menno I. Gaakeer, Gideon H.P. Latten, Jannet Mehagnoul, Özlem Tapirdamaz, Frits van Osch, Luc Mortelmans

## Abstract

**Objective:** Expanding staff levels is a strategy for hospitals to increase surge capacity. This study aimed to evaluate whether emergency healthcare workers (HCWs) are willing to work (WTW) during a crises or disaster and which working conditions would influence their decision.

**Methods:** HCWs of emergency departments (ED) and intensive care units (ICU) of five Dutch hospitals were surveyed about elevens disaster scenarios. For each scenario, HCWs were asked about their WTW and which conditions would influence their decision. Knowledge and perceived risk and danger was assessed per scenario.

**Results:** 306/630 HCWs completed the survey. An influenza epidemic, SARS-CoV-2 pandemic and natural disaster were associated with highest WTW rates (69.0%, 63.7% and 53.3% respectively). WTW was lowest in nuclear incident (4.6%) and dirty bomb (3.3%) scenarios. WTW was higher in physicians than in nurses. Male ED HCWS, single HCWs and childless HCWs were more often WTW. Personal protective equipment (PPE) and safety of HCWs’ family were the most important working conditions. Perceived knowledge scored lowest in dirty bomb, biological and nuclear incident scenarios. These scenarios rated highest with regards to perceived danger.

**Conclusions:** WTW depended on disaster type, profession and working department. Provision of PPE and safety of HCWs’ family were found to be predominant working conditions.

## INTRODUCTION

Mass casualty incidents (MCIs) are defined as ‘incidents leading to a surge of patients that overwhelm the local healthcare system’.^1^ MCIs can be natural disasters, such as floods or earthquakes, or man-made events, including infrastructural disasters or terrorist attacks. The past few decades have witnessed an increase in several hazards, including the risk of natural disasters and terrorist attacks^2, 3^, as well as an increase of internal hospital disasters.^4^ Combined internal and external events eventually affect the functioning of hospitals, acute care departments being particularly vulnerable.^4^ Not only disasters with a large number of patients affect hospital functioning. Treatment of specific types of patients, such as radiographic or biologically contaminated patients, can cause disruption. Furthermore, the COVID-19 pandemic demonstrated that large-scale infectious disease outbreaks pose a major burden on hospital functioning, with healthcare workers (HCWs) experiencing stress and moral injury, leading to increased burnout rates and staffing shortages.^5–7^ Crises and disasters have the potential to further exacerbate this burden and its associated risks.

To deal with MCIs, hospitals are equipped with disaster management plans. Such plans almost exclusively lean on the availability of additional staff. However, is it reasonable for hospitals to expect their attendance in all disaster scenarios? Previous studies focusing on the willingness to work (WTW) of emergency HCWs during a disaster showed that there were no scenarios in which all HCWs were WTW unconditionally.^8–10^ Only few studies have assessed the disaster preparedness of Dutch hospitals. Therefore, we aimed to investigate the WTW of Dutch emergency HCWs.

## METHODS

### STUDY DESIGN

A cross-sectional survey among ED and ICU HCWs in five Dutch hospitals.

### SETTING AND PARTICIPANTS

ED and ICU nurses and physicians of five Dutch hospitals (VieCuri Medical Centre, Venlo; Laurentius Hospital, Roermond; Zuyderland Medical Centre, Heerlen; Zuyderland Medical Centre, Sittard-Geleen; and Adrz Hospital, Goes) were invited to participate in this study. Data was collected through an online survey. All ages and gender groups were included. HCWs were informed via department management, email and posters. Information about the design and goal of the study was provided in the email containing the weblink to the survey. Informed consent was obtained through survey introduction, where HCWs were informed that if they continued the survey, they agreed with study participation. Participants were informed that their participation was voluntary and that study withdrawal was possible before and while filling in the questionnaire (by not completing it). After the questionnaire was completed, all results were fully anonymous and could not be traced back to the individual.

### SURVEY

The survey used, called Fight or Flight, was developed by the Centre for Research and Education in Emergency Care (CREEC) at the University of Leuven, Belgium. The survey comprised a minimum of 52 items, potentially extending to a maximum of 134, distributed in two sections: a demographic section consisting of nine questions and a scenario section consisting of eleven disaster scenarios with four questions each (willingness to come to work, perceived knowledge of disaster scenario, perceived risk and perceived danger of disaster scenario). When the first scenario question was answered with ‘willing to go to work under certain conditions’, an additional twelve statements with working conditions were presented. Participants had to rate the importance of these statements in association with willingness to come to work on a 10-point Likert scale. Knowledge of the scenario, perceived risk and perceived danger was also rated on a 10-point Likert scale.

The original survey was adjusted to fit the current time frame and situation in the Netherlands. Changes included replacing the Mexican fever and SARS pandemic outbreak scenarios for a renewed SARS-CoV-2 surge and a mass shooting scenario. Two versions of the survey were used - one for nurses and one for physicians, which only included differences in the demographic sections. See online supplemental 1 for an example of the survey.

### DATA COLLECTION AND ANALYSIS

Data collection took place from November 28th, 2022 until March 5th, 2023. The surveys were distributed and data was collected using Castor EDC (Amsterdam, the Netherlands); a cloud-based secured platform for electronic data collecting. All participants received a participant-ID when uploading their email-address into Castor. The email-address could not be traced back to answers by the researchers. Non-responders received a maximum of three e-mailed reminders. A general survey-link and QR-codes were distributed via e-mail and flyers to increase the response rate.

### STATISTICS

SPSS (IBM Corp. Released 2019. IBM SPSS Statistics for Windows, Version 26.0. Armonk, NY: IBM Corp) was used for data analyses. Data was presented descriptively as means and standard deviations (SDs) for normally distributed continuous data, as medians and interquartile ranges (IQR) for not normally distributed continuous data, and as frequencies and percentages for categorical data. Chi square tests were used for categorical demographic data. Fisher’s Exact tests were used for continuous demographic data and ordinal data.

The homogeneity of the items in the subscales of the Fight or Flight survey was analysed by calculating Cronbach’s Alpha using the Statistical Package for the Social Sciences (SPSS) version 20 (IBM, Texas, USA) in a previously performed study^8^. The Cronbach’s Alpha was 0.927 which is a satisfactory value.

### ETHICAL APPROVAL

A non-WMO declaration was granted by the medical ethical review board of Maastricht University Medical Centre (study ID 2022-3417).

## RESULTS

A total of 647 HCWs were invited to participate in the study. Of the 322 (50%) HCWs that completed the survey, 16 did not fit the job description and were subsequently excluded. Eventually, 306 participants were included in the analysis.

Most respondents were female (67.3%) and nurses represented 71.6% of participants. For 7 (3.2%) respondents the main department was unknown. Respondents were in a relationship in 85.3%, and 56.2% had children. About a third of physicians (32.2%) and 15.5% of nurses had any form of past disaster management training. Specific topic disaster training, such as disaster management, dealing with an epidemic/pandemic, chemical incidents, nuclear incidents or mass casualty incidents varied between 6.9% to 29.9% for physicians, and 5.5% to 18.7% for nurses (Table 1).

**TABLE 1:**
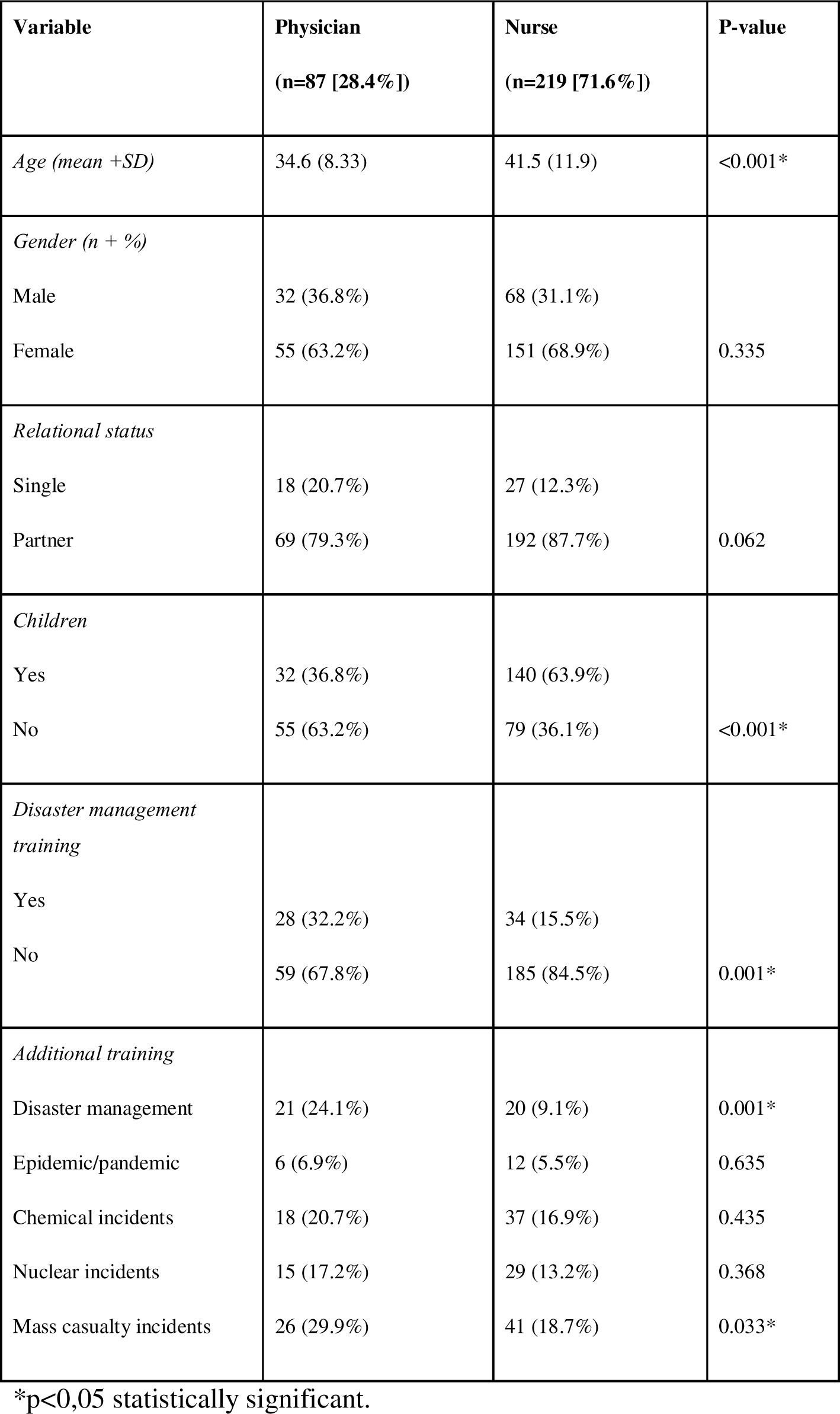
DEMOGRAPHIC DATA (N=306)

| **Variable** | **Physician**<br>**(n=87 [28.4%])** | **Nurse**<br>**(n=219 [71.6%])** | **P-value** |
| --- | --- | --- | --- |
| *Age (mean +SD)* | 34.6 (8.33) | 41.5 (11.9) | <0.001* |
| *Gender (n + %)* |  |  |  |
| Male | 32 (36.8%) | 68 (31.1%) | 0.335 |
| Female | 55 (63.2%) | 151 (68.9%) |  |
| *Relational status* |  |  |  |
| Single | 18 (20.7%) | 27 (12.3%) | 0.062 |
| Partner | 69 (79.3%) | 192 (87.7%) |  |
| *Children* |  |  |  |
| Yes | 32 (36.8%) | 140 (63.9%) | <0.001* |
| No | 55 (63.2%) | 79 (36.1%) |  |
| *Disaster management training* |  |  |  |
| Yes | 28 (32.2%) | 34 (15.5%) | 0.001* |
| No | 59 (67.8%) | 185 (84.5%) |  |
| *Additional training* |  |  |  |
| Disaster management | 21 (24.1%) | 20 (9.1%) | 0.001* |
| Epidemic/pandemic | 6 (6.9%) | 12 (5.5%) | 0.635 |
| Chemical incidents | 18 (20.7%) | 37 (16.9%) | 0.435 |
| Nuclear incidents | 15 (17.2%) | 29 (13.2%) | 0.368 |
| Mass casualty incidents | 26 (29.9%) | 41 (18.7%) | 0.033* |

\*p<0,05 statistically significant.

There was no disaster scenario in which all emergency HCWs were WTW unconditionally. HCWs were most likely to come to work during an influenza epidemic (69.0%), a SARS-CoV-2 pandemic (63.7%), or a natural disaster (53.3%). They were least likely to come during an ebola or haemorrhagic fever (HF) outbreak (31.4%), a nuclear incident (35.9%), or a dirty bomb (37.9%) (Figure 1).

**Figure 1.**
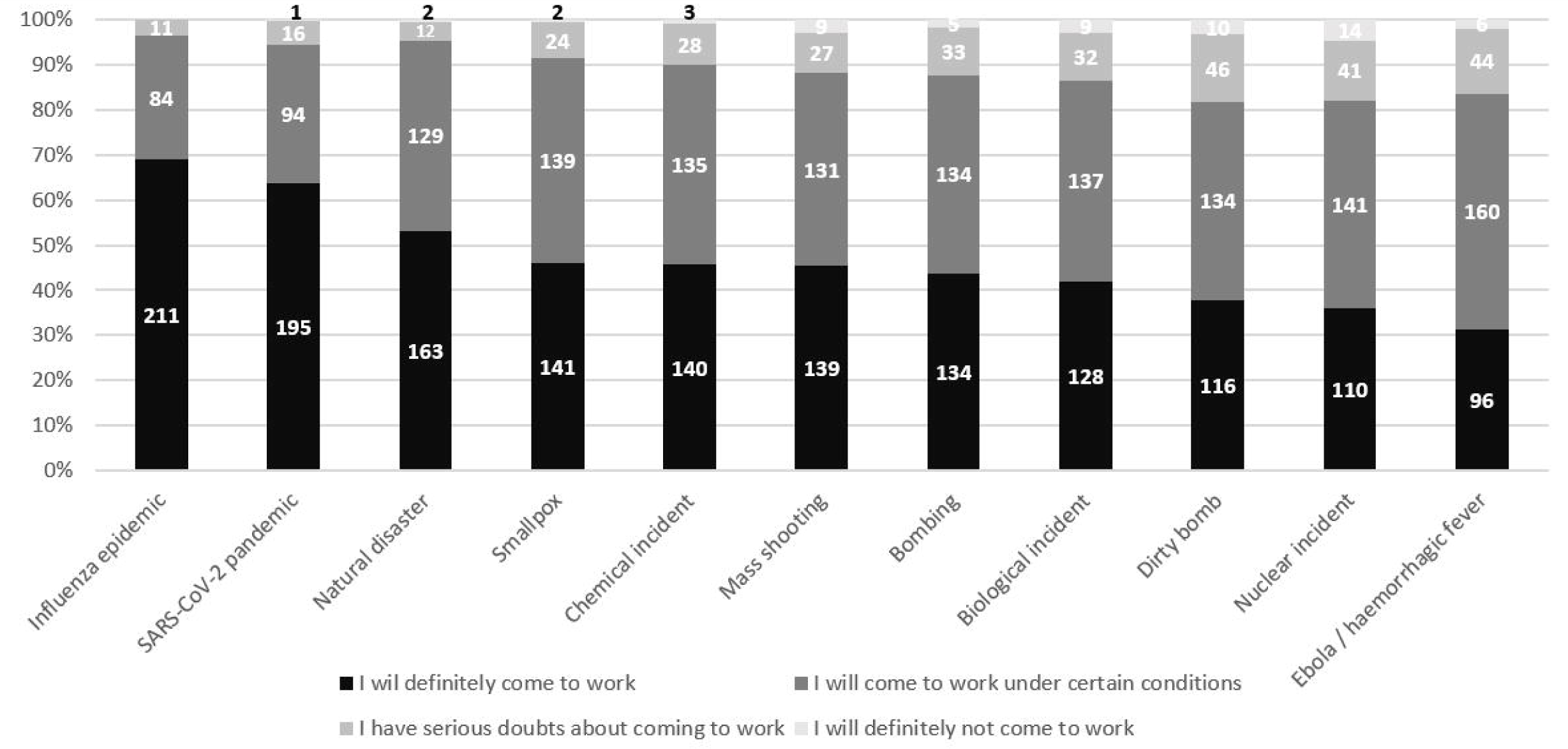
Willingness to work according to disaster scenario (n=306)

An influenza epidemic and a renewed SARS-CoV-2 pandemic were scenarios in which the lowest number of HCWs required working conditions (27.5% and 30.7% respectively). For all other scenarios, about half of all HCWs required working conditions (ranging from 42.2% during a natural disaster to 52.3% during an ebola or HF outbreak). In case of a dirty bomb, a nuclear incident, or an ebola or HF outbreak, the rates of HCWs that were unwilling to come to work or had serious doubts about coming to work were highest (18.3%, 18.0%, and 16.3% respectively). An influenza epidemic was the only disaster scenario in which no HCWs were unwilling to come to work.

Physicians were more WTW unconditionally during all disaster scenarios. The greatest differences were observed in the scenarios SARS-CoV-2 pandemic (physicians vs. nurse, 80.5% vs. 57.1%, p=<0.001) and chemical incident (physician vs. nurse, 62.1% vs. 39.3%, p=0.003). Nurses were more likely to set certain conditions to come to work than physicians, ranging from 30.6% during an influenza epidemic to 51.6% during an outbreak of ebola or HF. During the scenario of a dirty bomb, the highest number of nurses expressed doubt or unwillingness to come to work (20.1%). For physicians this was during the scenario of dirty bomb and nuclear incident (both 13.8%) (Table 2).

**TABLE 2:** WILLINGNESS TO WORK ACCORDING TO PROFESSION.

| Disaster scenario | I will definitely come to work |  | I will come to work under certain conditions |  | I have serious doubts about coming to work |  | I will definitely not come to work |  | Fisher's Exact test (p-value) |
| --- | --- | --- | --- | --- | --- | --- | --- | --- | --- |
|  | Physician (n=87) | Nurse (n=219) | Physician (n=87) | Nurse (n=219) | Physician (n=87) | Nurse (n=219) | Physician (n=87) | Nurse (n=219) |  |
| Natural disaster | 55 (63.2%) | 108 (49.3%) | 31 (35.6%) | 98 (44.7%) | - | 12 (5.5%) | 1 (1.1%) | 1 (0.5%) | 0,013* |
| Bomb attack | 49 (56.3%) | 85 (38.8%) | 34 (39.1%) | 100 (45.7%) | 4 (4.6%) | 29 (13.2%) | - | 5 (2.3%) | 0,010* |
| Influenza epidemic | 68 (78.2%) | 143 (65.3%) | 17 (19.5%) | 67 (30.6%) | 2 (2.3%) | 9 (4.1%) | - | - | 0,098 |
| SARS-CoV-2 pandemic | 70 (80.5%) | 125 (57.1%) | 15 (17.2%) | 79 (36.1%) | 2 (2.3%) | 14 (6.4%) | - | 1 (0.5%) | <0,001* |
| Mass shooting | 49 (56.3%) | 90 (41.1%) | 33 (37.9%) | 98 (44.7%) | 5 (5.7%) | 22 (10.0%) | - | 9 (4.1%) | 0,031* |
| Ebola/HF | 30 (34.5%) | 66 (30.1%) | 47 (54.0%) | 113 (51.6%) | 10 (11.5%) | 34 (15.5%) | - | 6 (2.7%) | 0,367 |
| Smallpox | 45 (51.7%) | 96 (43.8%) | 39 (44.8%) | 100 (45.7%) | 3 (3.4%) | 21 (9.6%) | - | 2 (0.9%) | 0,221 |
| Chemical incident | 54 (62.1%) | 86 (39.3%) | 28 (32.2%) | 107 (48.9%) | 5 (5.7%) | 23 (10.5%) | - | 3 (1.4%) | 0,003* |
| Biological incident | 39 (44.8%) | 89 (40.6%) | 40 (46.0%) | 97 (44.3%) | 6 (6.9%) | 26 (11.9%) | 2 (2.3%) | 7 (3.2%) | 0,615 |
| Nuclear incident | 35 (40.2%) | 75 (34.2%) | 40 (46.0%) | 101 (46.1%) | 8 (9.2%) | 33 (15.1%) | 4 (4.6%) | 10 (4.6%) | 0,531 |
| Dirty bomb | 42 (48.3%) | 74 (33.8%) | 33 (37.9%) | 101 (46.1%) | 11 (12.6%) | 35 (16.0%) | 1 (1.1%) | 9 (4.1%) | 0,099 |

\*p<0.05 statistically significant.

HCWs who had received previous disaster training were more WTW during all disaster scenarios. These differences were most pronounced in the disaster scenarios bomb attack (training vs. no training, 59.7% vs. 39.8%, p=0.030) and nuclear incident (training vs. no training, 51.6% vs. 32.0%, p=0.010). There were no HCWs with previous disaster training that were unwilling to come to work during a natural disaster, bomb attack, influenza epidemic, SARS-CoV-2 pandemic, mass shooting and chemical incident. For HCWs without previous disaster training this was only the case for an influenza epidemic (Table 3).

**TABLE 3:** WILLINGNESS TO WORK ACCORDING TO DISASTER TRAINING.

| Disaster scenario | I will definitely come to work |  | I will come to work under certain conditions |  | I have serious doubts about coming to work |  | I will definitely not come to work |  | Fisher's Exact test (p-value) |
| --- | --- | --- | --- | --- | --- | --- | --- | --- | --- |
| Disaster training | Yes (n=62) | No (n=244) | Yes (n=62) | No (n=244) | Yes (n=62) | No (n=244) | Yes (n=62) | No (n=244) |  |
| Natural disaster | 39 (62.9%) | 124 (50.8%) | 22 (35.5%) | 107 (43.9%) | 1 (1.6%) | 11 (4.5%) | - | 2 (0.8%) | 0,350 |
| Bomb attack | 37 (59.7%) | 97 (39.8%) | 22 (35.5%) | 112 (45.9%) | 3 (4.8%) | 30 (12.3%) | - | 5 (2.0%) | 0,030* |
| Influenza epidemic | 43 (69.4%) | 168 (68.9%) | 17 (27.4%) | 67 (27.5%) | 2 (3.2%) | 9 (3.7%) | - | - | 1 |
| SARS-CoV-2 pandemic | 43 (69.4%) | 152 (62.3%) | 15 (24.2%) | 79 (32.4%) | 4 (6.5%) | 12 (4.9%) | - | 1 (0.4%) | 0,535 |
| Mass shooting | 36 (58.1%) | 103 (42.2%) | 23 (37.1%) | 108 (44.3%) | 3 (4.8%) | 24 (9.8%) | - | 9 (3.7%) | 0,088 |
| Ebola/HF | 26 (41.9%) | 70 (28.7%) | 29 (46.8%) | 131 (53.7%) | 6 (9.7%) | 38 (15.6%) | 1 (1.6%) | 5 (2.0%) | 0,211 |
| Smallpox | 30 (48.8%) | 111 (45.5%) | 27 (43.5%) | 112 (45.9%) | 4 (6.5%) | 20 (8.2%) | 1 (1.6%) | 1 (0.4%) | 0,614 |
| Chemical incident | 36 (58.1%) | 104 (42.6%) | 23 (37.1%) | 112 (45.9%) | 3 (4.8%) | 25 (10.2%) | - | 3 (1.2%) | 0,153 |
| Biological incident | 28 (45.2%) | 100 (41.0%) | 29 (46.8%) | 108 (44.3%) | 4 (6.5%) | 28 (11.5%) | 1 (1.6%) | 8 (3.3%) | 0,685 |
| Nuclear incident | 32 (51.6%) | 78 (32.0%) | 26 (41.9%) | 115 (47.1%) | 3 (4.8%) | 38 (15.6%) | 1 (1.6%) | 13 (5.3%) | 0,010* |
| Dirty bomb | 31 (50.0%) | 85 (34.8%) | 26 (41.9%) | 108 (44.3%) | 4 (6.5%) | 42 (17.2%) | 1 (1.6%) | 9 (3.7%) | 0,054 |

\*p<0.05 statistically significant.

ED HCWs were more WTW unconditionally in all disaster scenario. Furthermore, ICU staff more often indicated that they were only WTW under certain conditions than ED staff in all scenarios. These rates were lowest for an influenza epidemic (35.4%) or a SARS-CoV-2 pandemic (38.8%). The most doubt or unwillingness to come to work was observed in the ebola/HF scenario (ED) and nuclear incident scenario (ICU) (Online supplemental 2).

When comparing WTW according to gender, significant differences were found for the scenarios bomb attack (male vs. female, 54.0% vs. 38.8%, p=0.037), ebola/HF (male vs. female, 39% vs. 27.7%, p=0.017) and dirty bomb (male vs. female, 39% vs. 37.4%, p=0.014) (Online supplemental 3).

Single HCWs demanded less working conditions, and there were less scenarios in which they would not show up at all. However, when comparing WTW according to relational status, no significant differences were found (Online supplemental 4).

When comparing WTW according to having children, significant differences were observed for chemical incident (children vs. no children, 39.0% vs. 54.5%, p=0.022) and biological incident (children vs. no children, 37.2% vs. 47.8%, p=0.025) (Online supplemental 5)

Some of the respondents indicated to be WTW under certain conditions. Having access to adequate PPE was the most important condition in every disaster scenario (median [interquartile range (IQR)]: 10 [8-10]). During a natural disaster, bombing, mass shooting, chemical incident, nuclear incident and dirty bomb, knowing that family is safe and taken care of was another condition deemed important (10 [8-10]). For the scenarios ebola or HF outbreak, smallpox outbreak, chemical incident, biological incident and nuclear incident being properly trained and kept updated on the development of the situation were the second-most important conditions to come to work. To be kept updated on the situation was also important during an influenza epidemic, a SARS-CoV-2 pandemic and a dirty bomb. Financial compensation was only deemed important in the case of an influenza epidemic (8 [6-10]) (Table 4).

**TABLE 4:** WILLINGNESS TO WORK UNDER CERTAIN CONDITIONS* MEDIAN + IQR.

**MEDIAN + IQR.**
| Disaster scenario | a | b | c | d | e | f | g | h |
| --- | --- | --- | --- | --- | --- | --- | --- | --- |
| **Natural disaster**<br>**n=129 (42,2%)** | 10<br>(8-10) | 9<br>(8-10) | 5<br>(2-8) | 6<br>(4-8) | 8<br>(7-9) | 10<br>(8-10) | 7<br>(5-9) | - |
| **Bombing n=134**<br>**(43.8%)** | 10<br>(8-10) | 9<br>(8-10) | 5,50<br>(3-8) | 8<br>(6-9) | 8<br>(7-10) | 10<br>(8-10) | 6,50<br>(5-9) | - |
| **Influenza pandemic**<br>**n=84 (27.5%)** | 8<br>(3-10) | 8<br>(4-10) | 6<br>(3-8) | 6,50<br>(5-8) | 8<br>(6,25-9) | 10<br>(9-10) | 8<br>(6-10) | 6<br>(3,25-8) |
| **SARS-CoV-2**<br>**pandemic n=94**<br>**(30.7%)** | 8<br>(2-9,25) | 8<br>(5-9) | 6,50<br>(2,75-8) | 8<br>(6-9) | 8<br>(7-10) | 10<br>(9-10) | 8<br>(6-10) | 6<br>(2-8) |
| **Mass-shooting**<br>**n=131 (42.8%)** | 10<br>(8-10) | 9<br>(8-10) | 5<br>(1-8) | 8<br>(6-9) | 8<br>(7-10) | 10<br>(8-10) | 7<br>(4-9) | - |
| **Ebola/HF n=160**<br>**(52.3%)** | 8<br>(4,25-10) | 8<br>(6-10) | 5<br>(1-8) | 9<br>(8-10) | 9<br>(8-10) | 10<br>(10-10) | 7<br>(5-10) | 8<br>(5-10) |
| **Smallpox n=139**<br>**(45.5%)** | 8<br>(5-10) | 8<br>(5-10) | 5<br>(1-8) | 8<br>(7-10) | 8<br>(7-10) | 10<br>(10-10) | 7<br>(5-10) | 8<br>(4-10) |
| **Chemical incident**<br>**n=135 (44.1%)** | 10<br>(8-10) | 9<br>(7-9) | 6<br>(2-8) | 9<br>(8-10) | 8<br>(8-10) | 10<br>(10-10) | 7<br>(5-10) | - |
| **Biological incident**<br>**n=137 (44.8%)** | 8<br>(4-10) | 8<br>(6-10) | 5<br>(1-8) | 9<br>(8-10) | 9<br>(8-10) | 10<br>(10-10) | 7<br>(5-9) | - |
| **Nuclear incident**<br>**n=141 (46.1%)** | 10<br>(8-10) | 9<br>(8-10) | 6<br>(2-8) | 9<br>(8-10) | 9<br>(8-10) | 10<br>(10-10) | 7<br>(5-9) | - |
| **Dirty bomb n=134**<br>**(43.8%)** | 10<br>(8-10) | 9<br>(8-10) | 5<br>(2-8) | 8<br>(7-10) | 9<br>(8-10) | 10<br>(10-10) | 7<br>(4-9) | - |

*Rated on a 10-point scale (10 = most important, 1 = least important).*
*\*(a) If I know my family is safe and taken care of; (b) If I am sure good communication lines with my family are available; (c) If my boss comes to work as well; (d) If I am trained to handle the situation; (e) If i get regular updates on the evolution of the incident; (f) If I am sure I can get adequate PPE; (g) If I get financial compensation; (h) I can get antivirals for free.*

Perceived knowledge was rated highest for an influenza epidemic (8 [7-8]) and SARS-CoV-2 pandemic (8 [8-9]). Lowest rates were reported for the scenarios dirty bomb, biological incident, nuclear incident and ebola or HF outbreak (3 [2-5], 4 [2-5], 4 [2-5], and 4 [3-5] respectively). Perceived risk was highest for an influenza epidemic (8 [7-9]) and SARS-CoV-2 pandemic (7 [5-8]). This was lowest for an ebola or HF outbreak and dirty bomb (both 3 [2-5]). Perceived danger was highest for a nuclear incident and dirty bomb (both 8 [7-9]), as well as for an ebola or HF outbreak (8 [6-9]) and a mass shooting (8 [5-9]). Perceived danger was lowest for a natural disaster, an influenza epidemic and a smallpox outbreak (5 [5-7] all) (Online supplemental 6).

## STRENGTHS AND LIMITATIONS

To our knowledge, a survey-based WTW study had not been performed among Dutch emergency HCWs yet. Strengths of our study include the multicentre design and specific focus on emergency healthcare, as these are a hospitals’ first line of response during a crises or disaster.

The total estimated response rate was 49%, despite placing maximum effort on inclusion. Survey-based studies performed within the ED or ICU generally show response-rates varying from 21.8% to 51%^7, 11–15^, which implies that the response to the present survey is satisfactory. Unfortunately, we were not able to asses non-response bias due to the anonymity of the study. However, it is known that survey studies with a smaller sample size (<500 participants) need a response rate of 20-25% to provide fairly confident estimates.^16^ Higher response rates are known to only cause minimal – or even non-existent – differences in estimated outcomes.^16^

The survey was considered to be extensive by some participants, which may be the reason 85 participants started but did not complete it.

The use of a 10-point Likert scale can be seen as a limitation, with it being unclear what the difference between scoring a certain number entails. The ‘estimation of risk or danger’ may have been multi-interpretable. Does the risk or danger entail the respondent or its environment?

In the web-based survey, hypothetical disaster scenarios were presented to the participants. They might not necessarily provide an accurate representation of the WTW during an actual emergency. Most of the previous research done in this field is based on hypothetical scenarios.

## DISCUSSION

This study assessed the willingness of Dutch emergency HCWs to come to work during crises or disasters. HCWs were most likely to respond unconditionally during an influenza epidemic, a SARS-CoV-2 surge or natural disaster. The scenarios nuclear or biological incident, dirty bomb and an ebola or HF outbreak were most often associated with doubts among HCWs. In the event of a disaster, emergency HCWs are more likely to respond to hospitals’ efforts to increase surge capacity if they have access to adequate PPE. The assurance that family is safe and having adequate training were other important conditions. Low perceived knowledge on certain (chemical – biological – radiographic – nuclear (CBRN)) disaster scenarios was associated with a lower willingness to come to work.

Previous studies also showed that there is no disaster scenario in which all HCWs are WTW unconditionally. Willingness to work depends on the type of disaster.^9, 10^ A similar study performed in Saudi-Arabia, showed the highest WTW during a natural disaster (62%), and a seasonal influenza pandemic (53%).^7^ The high willingness to respond to natural disasters resonates with previous research too.^9, 10, 17^ Influenza epidemics are a regular occurrence in the Netherlands and the high WTW may be explained by the high perceived level of knowledge and experience, and therefore feeling prepared. The same may hold true for a new SARS-CoV-2 surge. WTW during the COVID-19 pandemic was previously shown to be high among physicians and nurses across the world.^18–20^ Important conditions that were set included higher salary (78.5%), better working conditions (72.1%), sufficient PPE (54.3%) and further education/training (62%).^20^

Similar to our study, an ebola outbreak emerged in Saudi Arabia and Belgium as one of the disasters for which respondents expressed the most serious doubts about coming to work.^8, 21^ Contrastingly, a study performed in Nigeria where a hypothetical Ebola virus outbreak was presented, showed that the majority of staff was WTW during an outbreak (73.1%).^22^ This difference may be explained by the fact that Nigeria has experienced the West African ebola epidemic in 2014-2016 and is one of the African countries which has seen the highest number of ebola cases.^23^ This suggests that perceived knowledge and experience of certain disaster scenarios influences the WTW.

In the case of a CBRN event, levels of WTW are usually lower than in other disaster scenarios. HCWs were less likely to respond to a CBRN event if they felt less prepared for such a scenario.^10, 17, 21, 24, 25^ This can be attributed to a lack of formal education and awareness of radiation-related events.^25^ Exposure risk and the lack of training in PPE increased anxiety among staff members, causing hesitancy among HCW to treat radiographically exposed patients.^27, 28^ Adequate training in PPE application was found to be a core competency in a review of disaster nursing practices.^29^ This is in line with recommendations by the World Health Organization and the International Council of Nursing (ICN) who advocate for training and rehearsals of disaster scenarios and PPE use.^30^

Most previous studies observed that female HCWs are less likely to come to work unconditionally than their male counterparts, as can be seen in our study during the scenarios bomb attack, ebola/HF and dirty bomb.^11, 17, 25, 31, 32^ This difference was attributed to traditional role patterns in which women are most responsible for the care of their children. HCWs with more family responsibilities, especially small children and pets, show less WTW in an emergency context.^10, 17, 31, 33^ This phenomenon was corroborated by the findings of our study.

Unwillingness to come to work in case of an infectious disease outbreak was mainly based on the fear of contracting an infectious disease and spreading it to family members ^9, 10, 28, 34^, perhaps explaining why single HCWs were more WTW during such scenarios.

Safe shelter for family and kids was an important factor in the decision to come to work during a disaster^10, 21^, as well as being able to communicate with family members while at work.^9^ In this study, knowing family is safe and taken care of was most important in the case of a natural disaster, bombing or mass-shooting.

## CONCLUSIONS

In this study, willingness to work depended on the type of disaster, the profession and department of the HCW. Access to adequate PPE was found to be the most important condition, followed by the assurance that HCWs family is safe. HCWs with a lower perceived knowledge about a certain (CBRN) disaster scenario were less likely to respond to work during a disaster. Hospitals should provide adequate education and training of different crises and disaster.

## Data Availability

All data produced in the present study are available upon reasonable request to the authors

## ACKNOWLEDGEMENTS

All authors had an equal contribution to creating this concept article for conceptualization and for writing the manuscript. All authors read and approved the final manuscript.

## AUTHOR CONTRIBUTIONS

LM conceived and designed the original study and survey. LMJE and DGB set up the study as described in this article. FHMvO provided advice on the study design. LE, DGB, TJTB, MIG, GHPL, JM and ÖT undertook the recruitment of participants in each corresponding hospital. LE managed the data and performed data analysis. DGB and FHMvO supervised and aided in the data analysis. LE drafted the manuscript. All authors contributed substantially to its revision. DGB takes responsibility for the paper as a whole.

All authors attest to meeting the four ICMJE.org authorship criteria: 1) Substantial contributions to the conception or design of the work; or the acquisition, analysis, or interpretation of data for the work; AND 2) Drafting the work or revising it critically for important intellectual content; AND 3) Final approval of the version to be published; AND 4) Agreement to be accountable for all aspects of the work in ensuring that questions related to the accuracy or integrity of any part of the work are appropriately investigated and resolved.

## LIST OF ABBREVIATIONS

CBRN: chemical, biological, radiographic, nuclear
CREEC: Centre for Research and Education in Emergency Care
ED: emergency department
HCW: healthcare workers
HF: haemorrhagic fever
ICN: International Council of Nursing
ICU: intensive care department
IQR: interquartile range
MCI: mass casualty incident
PPE: personal protective equipment
SD: standard deviation
SPSS: Statistical Package for the Social Sciences
WTW: willingness to work

Online supplemental 1 – Fight or Flight survey

Online supplemental 2 – Willingness to work according to department

Online supplemental 3 – willingness to work according to gender

Online supplemental 4 – Willingness to work according to relational status

Online supplemental 5 – Willingness to work according to having children

Online supplemental 6 – Participants’ self-ratings on their perceived knowledge, risk and danger of disasters

## REFERENCES

1. World Health Organization. Mass Casualty Management Systems: Strategies and Guidelines for Building Health Sector Capacity. World Health Organization. 2007. https://apps.who.int/iris/bitstream/handle/10665/43804/9789241596053_eng.pdf

2. Ritchie H, Rosado P, Roser M. Natural Disasters. Our World In Data. 2022 https://ourworldindata.org/natural-disasters

3. Centre for Research on the Epidemiology of Disasters (CRED), UN Office for Disaster Risk Reduction (UNDRR). Human cost of disasters: An overview of the last 20 years, 2000-2019. 2020 https://www.undrr.org/publication/human-cost-disasters-overview-last-20-years-2000-2019

4. Klokman V, Barten D, Peters N, et al. A scoping review of internal hospital crises and disasters in the Netherlands, 2000-2020. PloS One. 2021 https://journals.plos.org/plosone/article?id=10.1371/journal.pone.0250551

5. Vizheh M, Qorbani M, Arzaghi S, et al. The mental health of healthcare workers in the COVID-19 pandemic: a systematic review. J Diabetes Metab Disord. 2020 https://link.springer.com/article/10.1007/s40200-020-00643-9

6. Shaukat N, Ali D, Razzak J. Physical and mental health impacts of COVID-19 on healthcare workers: a scoping review. Int J Emerg Med. 2020 https://intjem.biomedcentral.com/articles/10.1186/s12245-020-00299-5

7. Hesselink G, Straten L, Gallée L, et al. Holding the frontline: a cross-sectional survey of emergency department staff well-being and psychological distress in the course of the COVID-19 outbreak. BMC Health Serv Res. 2021 https://bmchealthservres.biomedcentral.com/articles/10.1186/s12913-021-06555-5#ref-CR7

8. Sultan MAS, Løwe Sørensen J, Carlström E, et al. Emergency healthcare providers’ perceptions of preparedness and willingness to work during disasters and public health emergencies. Healthcare (Basel). 2020 https://pubmed.ncbi.nlm.nih.gov/33138164/

9. Arbon P, Cusack L, Ranse J, et al. Exploring staff willingness to attend work during a disaster: A study of nurses employed in four Australian emergency departments. Aust Emerg Nurs J. 2012 https://www.sciencedirect.com/science/article/abs/pii/S1574626713000360

10. Chaffee M. Willingness of health care personnel to work in a disaster: an integrative review of the literature. Disaster Med Public Health Prep. 2009 https://pubmed.ncbi.nlm.nih.gov/19293743/

11. Oerlemans A, Wollersheim H, van Sluisveld N, et al. Rationing in the intensive care unit in case of full bed occupancy: a survey among intensive care unit physicians. BMC Anesthesiology. 2016 https://bmcanesthesiol.biomedcentral.com/articles/10.1186/s12871-016-0190-5

12. Hickey M, Yadav K, Abdulaziz K, et al. Attitudes and acceptability of organ and tissue donation registration in the emergency department: a national survey of emergency physicians. CJEM. 2022 https://pubmed.ncbi.nlm.nih.gov/35124786/

13. Edwards M, Naik P, Bachuwa G, et al. Survey of Emergency Department Staff About Challenges and Recommendations for Emergency Department Care of Extended Care Facility Patients. JAMDA. 2012 https://pubmed.ncbi.nlm.nih.gov/21450188/

14. Sklar D, Crandall C, Zola T, et al. Emergency Physician Perceptions of Patient Safety Risks. Ann Emerg Med. 2009 https://www.annemergmed.com/article/S0196-0644(09)01447-4/fulltext

15. Opgenorth D, Stelfox H, Gilfoyle E, et al. Perspectives on strained intensive care unit capacity: A survey of critical care professionals. PLoS One. 2018 https://pubmed.ncbi.nlm.nih.gov/30133479/

16. Fosnacht K, Sarraf S, Howe E, et al. How important are high response rates for college surveys? Rev High Ed. 2017 https://muse.jhu.edu/article/640611

17. Brice J, Gregg D, Sawyer D, et al. Survey of hospital employees’ personal preparedness and willingness to work following a disaster. South Med J. 2017 https://sma.org/southern-medical-journal/article/survey-hospital-employees-personal-preparedness-willingness-work-following-disaster/

18. Rafi A, Hasan T, Tasnia Azad D, et al. Willingness to work during initial lockdown due to COVID-19 pandemic: study based on an online survey among physicians of Bangladesh. PloS One. 2021 https://journals.plos.org/plosone/article?id=10.1371/journal.pone.0245885

19. Lord H, Loveday C, Moxham I, et al. Effective communication is key to intensive care nurses’ willingness to provide nursing care amidst the COVID-19 pandemic. Intensive Crit Care Nurs. 2021 https://www.ncbi.nlm.nih.gov/pmc/articles/PMC7528824/

20. Hoffmann Kusk K, Laugesen B, Germund Nielsen M. Willingness and preparedness to work during the first wave of the COVID-19 pandemic: a cross-sectional survey among registered nurses in a Danisch university hospital. Nord. J. Nurs. Res. 2023 https://journals.sagepub.com/doi/10.1177/20571585221150225

21. Hendrickx C, Van Turnhout P, Mortelmans L, et al. Willingness to Work of Hospital Staff in Disasters: A Pilot Study in Belgian Hospitals. Prehosp. Disaster Med. 2017 https://www.researchgate.net/publication/311935532_Willingness_to_work_of_hospital_staff_in_disasters_A_pilot_study_in_Belgian_hospitals

22. Ibiok N, Onyedinma C, Agwu-Umahi O, et al. Healthcare Workers’ Willingness to Report to Work During a Pandemic in Southeastern Nigeria: A Hypothetical Case Using Ebola Virus Disease. Int J Med Health Dev. 2022 https://www.ijmhdev.com/showBackIssue.asp?issn=2635-3695;year=2023;volume=28;issue=1;month=January-March

23. Centres for Disease Control and Prevention. History of Ebola Outbreaks. Last reviewed January 27 2023 https://www.cdc.gov/vhf/ebola/history/chronology.html

24. O’Sullivan T, Dow D, Turner M, et al. Disaster and emergency management: Canadian nurses’ perceptions of preparedness on hospital front lines. Prehosp Disaster Med. 2008 https://pubmed.ncbi.nlm.nih.gov/18702283/

25. Santinha G, Forte T, Gomes A. Willingness to Work during Public Health Emergencies: A Systematic Literature Review. Healthc (Amst). 2022 https://www.mdpi.com/2227-9032/10/8/1500

26. Shimada J, Tase C, Tsukada Y, et al. Early-stage responses of intensive care units during major disasters: from the experiences of the great East Japan earthquake. J Med Sci. 2015 https://www.jstage.jst.go.jp/article/fms/61/1/61_2014-35/_article

27. Balicer R, Catlett C, Barnett D, et al. Characterizing hospital workers’ willingness to respond to a radiological event. PloS One. 2011 https://journals.plos.org/plosone/article?id=10.1371/journal.pone.0025327

28. Sellers D, Ranse J. The impact of mass casualty incidents on intensive care units. Aus Crit Care. 2019 https://pubmed.ncbi.nlm.nih.gov/31980255/

29. Thobaity A, Plummer V, Williams B. What are the most common domains of the core competencies of disaster nursing? A scoping review. Int Emerg Nurs. 2017 https://www.sciencedirect.com/science/article/pii/S1755599X1630163X

30. World Health Organization, International Council of Nurses. ICN framework of disaster nursing competencies. 2009 https://www.who.int/Westernpacific?ua=1

31. Aoyagi Y, Beck C, Dingwall R, et al. Healthcare workers’ willingness to work during an influenza pandemic: a systematic review and meta-analysis. Influenza Other Respir Viruses. 2015 https://onlinelibrary.wiley.com/doi/10.1111/irv.12310

32. Al-Hunaishi W, Hoe V, Chinna K. Factors associated with healthcare workers willingness to participate in disasters: A cross-sectional study in Sana’a, Yemen. BMJ Open. 2019. https://bmjopen.bmj.com/content/9/10/e030547

33. Hill M, Smith E, Mills B. Willingness to work amongst Australian frontline healthcare workers during Australia’s first wave of COVID-19 community transmission: results of an online survey. Disaster Med. Public Health Prep. 2021 https://www.ncbi.nlm.nih.gov/pmc/articles/PMC8545838/pdf/S1935789321002883a.pdf

34. Morley C, Unwin M, Peterson GM, et al. Emergency department crowding: A systematic review of causes, consequences and solutions. PLoS One. 2018 https://pubmed.ncbi.nlm.nih.gov/30161242/

