## Appendix I for "Fight or Flight: Emergency Healthcare Workers’ Willingness to Work during Crises and Disasters: A cross-sectional multicentre study in the Netherlands"

**ONLINE SUPPLEMENTALS**

**ONLINE SUPPLEMENTAL 1: SURVEY FIGHT OR FLIGHT**

*Translated to English for the purposes of publication. Dutch survey used in study.*

Part 1: Demographic data

1) What is your gender?
   a) Male
   b) Female
2) At which hospital do you work?
   a) VieCuri Medical Centre, Venlo
   b) Laurentius Hospital, Roermond
   c) Zuyderland Medical Centre, Heerlen/Sittard
   d) Admiraal de Ruyter Hospital, Goes
3) What is your age (in years)?
4) What is your relationship status?
   a) Single
   b) Partner
5) Do you have children?
   a) Yes
   b) No
6) If yes, do your children live with you?
   a) Yes
   b) No
7) Have you had previous disaster training?
   a) Yes
   b) No
8) What is your medical profession?

*Survey physician*

a) Emergency doctor
b) Intensivist
c) Other medical specialty
d) Physician assistant emergency department
e) Doctor in training to emergency doctor
f) Doctor not in training to emergency doctor
g) Physician assistant intensive care unit
h) Doctor in training to intensivist
i) Doctor not in training to intensivist
j) Doctor in training other medical specialty
k) Doctor not in training other medical specialty
l) Other

*Survey nurse*

m) MBO nurse
n) HBO nurse
o) Basic acute care nurse
p) Emergency department nurse
q) Intensive care unit nurse
r) Emergency department + intensive care unit nurse
s) Emergency department nurse in training
t) Intensive care unit nurse in training
u) Plaster technician
v) Doctor’s assistant emergency department
w) Doctor’s assistant intensive care unit
x) Bachelor medical assistance
y) Other

9) Have you had any specific additional training?
   a) Disaster management/disaster medicine
   b) Dealing with an epidemic/pandemic
   c) Chemical incidents
   d) Nuclear incidents
   e) Mass casualty incidents

Part 2: disaster scenario’s

1) Natural disaster close to home and/or work (eg. flood)
2) Severe bombing (eg. London or Madrid bombing)
3) Severe, but classic flu epidemic
4) Severe revival SARS-CoV-2 pandemic
5) Mass-shooting with war weapons (eg. Bataclan (Paris) shooting)
6) Outbreak ebola/haemorrhagic fever
7) Outbreak smallpox
8) Chemical incident (industrial, transport, terrorist attack)
9) Biological incident (eg. Anthrax)
10) Nuclear incident (eg. leakage nuclear power plant, accident isotopetransport)
11) Dirty bomb

Example of questions asked per disaster scenario:

Here we would like to evaluate the impact of [disaster scenario] in the near proximity of your hospital/where possibly infected or contaminated patients will be transported to your hospital. How do you think you would react if such a scenario presents itself in your (working) environment?

1) How do you see working in these circumstances?
   a) I am definitely coming to work
   b) I am coming to work under certain conditions
   c) I have serious doubts about coming to work
   d) I am definitely not coming to work (even if it costs me my job)
2) If you have answered question 1 with ‘willing to come to work under certain working conditions’, which conditions may convince you to come to work? U can choose multiple answers, but you do not have to. Rate your answers of importance (10 = most important, 1 = least important)
   a) If I know there is safe shelter for my family/children organised

(i) 10 – 9 – 8 – 7 – 6 – 5 – 4 – 3 – 2 – 1 – not applicable

b) If there are adequate communication lines organised with family/children (phone, internet)
c) If my boss comes to work
d) If I had adequate explanation/training beforehand for this situation
e) If I am kept up to date on the situation
f) If I am sure I get adequate personal protective equipment
g) If I get a financial compensation
h) If I get free special medication (eg. Tamiflu) (*for scenario flu epidemic, SARS-CoV-1 pandemic, outbreak ebola/HF, outbreak smallpox*)

3) How do you further think of this situation? Rated on a scale from 1-10 (1 = lowest, 10 = highest):
   a) My knowledge of this situation is

(i) 10 – 9 – 8 – 7 – 6 – 5 – 4 – 3 – 2 – 1

b) I estimate the risk of this scenario happening as
c) I estimate the danger associated with this scenario as

**ONLINE SUPPLEMENTAL 2: WILLINGNESS TO WORK ACCORDING TO DEPARTMENT**

| **Disaster scenario** | **I will definitely come to work** |  | **I will come to work under certain conditions** | |  | **I have serious doubts about coming to work** |  | **I will definitely not come to work** |  | **Fisher’s Exact test (p-value)** |
| --- | --- | --- | --- | --- | --- | --- | --- | --- | --- | --- |
|  | **ED (n=152)** | **ICU (n=147)** | **ED (n=152)** | **ICU (n=147)** | | **ED (n=152)** | **ICU (n=147)** | **ED (n=152)** | **ICU (n=147)** |  |
| **Natural disaster** | 93 (61.2%) | 66 (44.9%) | 53 (34.9%) | 73 (49.7%) | | 5 (3.3%) | 7 (4.8%) | 1 (0.7%) | 1 (0.7%) | 0,125 |
| **Bomb attack** | 76 (50.0%) | 55 (37.4%) | 64 (42.1%) | 67 (45.6%) | | 9 (5.9%) | 24 (16.3%) | 3 (2.0%) | 1 (0.7%) | 0,009* |
| **Influenza epidemic** | 115 (75.7%) | 91 (61.9%) | 30 (19.7%) | 52 (35.4%) | | 7 (4.6%) | 4 (2.7%) | - | - | 0,036* |
| **SARS-CoV-2 pandemic** | 106 (69.7%) | 84 (57.1%) | 35 (23.0%) | 57 (38.8%) | | 11 (7.2%) | 5 (3.4%) | - | 1 (0.7%) | 0,038* |
| **Mass shooting** | 78 (51.3%) | 57 (38.8%) | 60 (39.5%) | 69 (46.9%) | | 11 (7.2%) | 16 (10.9%) | 3 (2.0%) | 5 (3.4%) | 0,151 |
| **Ebola/HF** | 52 (34.2%) | 42 (28.6%) | 77 (50.7%) | 79 (53.7%) | | 19 (12.5%) | 24 (16.3%) | 4 (2.6%) | 2 (1.4%) | 0,817 |
| **Smallpox** | 76 (50.0%) | 62 (42.2%) | 65 (42.8%) | 71 (48.3%) | | 10 (6.6%) | 13 (8.8%) | 1 (0.7%) | 1 (0.7%) | 0,681 |
| **Chemical incident** | 79 (52.0%) | 59 (40.1%) | 60 (39.5%) | 71 (48.3%) | | 11 (7.2%) | 16 (10.9%) | 2 (1.3%) | 1 (0.7%) | 0,271 |
| **Biological incident** | 70 (46.1%) | 56 (38.1%) | 67 (44.1%) | 66 (44.9%) | | 11 (7.2%) | 20 (13.6%) | 4 (2.6%) | 5 (3.4%) | 0,442 |
| **Nuclear incident** | 66 (43.4%) | 41 (27.9%) | 66 (43.4%) | 72 (49.0%) | | 14 (9.2%) | 26 (17.7%) | 6 (3.9%) | 8 (5.4%) | 0,081 |
| **Dirty bomb** | 66 (43.4%) | 48 (32.7%) | 65 (42.8%) | 66 (44.9%) | | 19 (12.5%) | 26 (17.7%) | 2 (1.3%) | 7 (4.8%) | 0,117 |

*p<0.05 statistically significant.

**ONLINE SUPPLEMENTAL 3: WILLINGNESS TO WORK ACCORDING TO GENDER**

| **Disaster scenario** | **I will definitely come to work** |  | **I will come to work under certain conditions** |  | **I have serious doubts about coming to work** |  | **I will definitely not come to work** |  | **Fisher’s Exact test (p-avlue)** |
| --- | --- | --- | --- | --- | --- | --- | --- | --- | --- |
|  | **Male (n=100)** | **Female (n=206)** | **Male** | **Female** | **Male** | **Female** | **Male** | **Female** |  |
| **Natural disaster** | 59 (59.0%) | 104 (50.5%) | 35 (35.0%) | 94 (45.6%) | 4 (4.0%) | 8 (3.9%) | 2 (2.0%) | - | 0,079 |
| **Bomb attack** | 54 (54.0%) | 80 (38.8%) | 38 (38.0%) | 96 (46.6%) | 6 (6.0%) | 27 (13.1%) | 2 (2.0%) | 3 (1.5%) | 0,037* |
| **Influenza epidemic** | 66 (66.0%) | 145 (70.4%) | 29 (29.0%) | 55 (26.7%) | 5 (5.0%) | 6 (2.9%) | - | - | 0,495 |
| **SARS-CoV-2 pandemic** | 66 (66.0%) | 129 (62.6%) | 28 (28.0%) | 66 (32.0%) | 6 (6.0%) | 10 (4.9%) | - | 1 (0.5%) | 0,804 |
| **Mass shooting** | 52 (52.0%) | 87 (42.2%) | 38 (38.0%) | 93 (45.1%) | 7 (7.0%) | 20 (9.7%) | 3 (3.0%) | 6 (2.9%) | 0,425 |
| **Ebola/HF** | 39 (39.0%) | 57 (27.7%) | 41 (41.0%) | 119 (57.8%) | 16 (16.0%) | 28 (13.6%) | 4 (4.0%) | 2 (1.0%) | 0,017* |
| **Smallpox** | 49 (49.0%) | 92 (44.7%) | 42 (42.0%) | 97 (47.1%) | 7 (7.0%) | 17 (8.3%) | 2 (2.0%) | - | 0,213 |
| **Chemical incident** | 52 (52.0%) | 88 (42.7%) | 38 (38.0%) | 97 (47.1%) | 8 (8.0%) | 20 (9.7%) | 2 (2.0%) | 1 (0.5%) | 0,205 |
| **Biological incident** | 41 (41.0%) | 87 (42.2%) | 42 (42.0%) | 95 (46.1%) | 11 (11.0%) | 21 (10.2%) | 6 (6.0%) | 3 (1.5%) | 0,194 |
| **Nuclear incident** | 42 (42.0%) | 68 (33.0%) | 37 (37.0%) | 104 (50.5%) | 15 (15.0%) | 26 (12.6%) | 6 (6.0%) | 8 (3.9%) | 0,149 |
| **Dirty bomb** | 39 (39.0%) | 77 (37.4%) | 41 (41.0%) | 93 (45.1%) | 12 (12.0%) | 34 (16.5%) | 8 (8.0%) | 2 (1.0%) | 0,014* |

*p<0,05 statistically significant.

**ONLINE SUPPLEMENTAL 4: WILLINGNESS TO WORK ACCORDING TO RELATIONAL STATUS**

| **Disaster scenario** | **I will definitely come to work** |  | **I will come to work under certain conditions** |  | **I have serious doubts about coming to work** |  | **I will definitely not come to work** |  | **Fisher’s Exact test (p-value)** |
| --- | --- | --- | --- | --- | --- | --- | --- | --- | --- |
|  | **Single (n=45)** | **Partner (n=261)** | **Single** | **Partner** | **Single** | **Partner** | **Single** | **Partner** |  |
| **Natural disaster** | 31 (68.9%) | 132 (50.6%) | 13 (28.9%) | 116 (44.4%) | 1 (2.2%) | 11 (4.2%) | - | 2 (0.8%) | 0,139 |
| **Bomb attack** | 24 (53.3%) | 110 (42.1%) | 15 (33.3%) | 119 (45.6%) | 5 (11.1%) | 28 (10.7%) | 1 (2.2%) | 4 (1.5%) | 0,362 |
| **Influenza epidemic** | 35 (77.8%) | 176 (67.4%) | 8 (17.8%) | 76 (29.1%) | 2 (4.4%) | 9 (3.4%) | - | - | 0,253 |
| **SARS-CoV-2 pandemic** | 32 (71.1%) | 163 (62.5%) | 11 (24.4%) | 83 (31.8%) | 2 (4.4%) | 14 (5.4%) | - | 1 (0.4%) | 0,652 |
| **Mass shooting** | 24 (53.3%) | 115 (44.1%) | 17 (37.8%) | 114 (43.7%) | 4 (8.9%) | 23 (8.8%) | - | 9 (3.4%) | 0,559 |
| **Ebola/HF** | 19 (42.2%) | 77 (29.5%) | 22 (48.9%) | 138 (52.9%) | 3 (6.7%) | 41 (15.7%) | 1 (2.2%) | 5 (1.9%) | 0,196 |
| **Smallpox** | 27 (60.0%) | 114 (43.7%) | 15 (33.3%) | 124 (47.5%) | 2 (4.4%) | 22 (8.4%) | 1 (2.2%) | 1 (0.4%) | 0,076 |
| **Chemical incident** | 29 (64.4%) | 111 (42.5%) | 14 (31.1%) | 121 (46.4%) | 2 (4.4%) | 26 (10.0%) | - | 3 (1.1%) | 0,062 |
| **Biological incident** | 25 (55.6%) | 103 (39.5%) | 17 (37.8%) | 120 (46.0%) | 2 (4.4%) | 30 (11.5%) | 1 (2.2%) | 8 (3.1%) | 0,192 |
| **Nuclear incident** | 23 (51.1%) | 87 (33.3%) | 19 (42.2%) | 122 (46.7%) | 2 (4.4%) | 39 (14.9%) | 1 (2.2%) | 13 (5.0%) | 0,065 |
| **Dirty bomb** | 25 (55.6%) | 91 (34.9%) | 15 (33.3%) | 119 (45.6%) | 4 (8.9%) | 42 (16.1%) | 1 (2.2%) | 9 (3.4%) | 0,075 |

*p<0,05 statistically significant.

**ONLINE SUPPLEMENTAL 5: WILLINGNESS TO WORK ACCORDING TO HAVING CHILDREN**

| **Disaster scenario** | **I will definitely come to work** |  | **I will come to work under certain conditions** |  | **I have serious doubts about coming to work** |  | **I will definitely not come to work** |  | **Fisher’s Exact test (p-value)** |
| --- | --- | --- | --- | --- | --- | --- | --- | --- | --- |
|  | **Children(n=172)** | **No children (n=134)** | **Children** | **No children** | **Children** | **No children** | **Children** | **No children** |  |
| **Natural disaster** | 89 (51.7% | 74 (55.2%) | 75 (43.6%) | 54 (40.3%) | 6 (3.5%) | 6 (4.5%) | 2 (1.2%) | - | 0,656 |
| **Bomb attack** | 75 (43.6%) | 59 (44.0%) | 76 (44.2%) | 58 (43.3%) | 19 (11.0%) | 14 (10.4%) | 2 (1.2%) | 3 (2.2%) | 0,921 |
| **Influenza epidemic** | 116 (67.4%) | 95 (70.9%) | 52 (30.2%) | 32 (23.9%) | 4 (2.3%) | 7 (5.2%) | - | - | 0,241 |
| **SARS-CoV-2**  **pandemic** | 108 (62.8%) | 87 (64.9%) | 55 (32.0%) | 39 (29.1%) | 9 (5.2%) | 7 (5.2%) | - | 1 (0.7%) | 0,791 |
| **Mass shooting** | 75 (43.6%) | 64 (47.8%) | 77 (44.8%) | 54 (40.3%) | 14 (8.1%) | 13 (9.7%) | 6 (3.5%) | 3 (2.2%) | 0,746 |
| **Ebola/HF** | 51 (29.7%) | 45 (33.6%) | 92 (53.5%) | 68 (50.7%) | 25 (14.5%) | 19 (14.2%) | 4 (2.3%) | 2 (1.5%) | 0,860 |
| **Smallpox** | 73 (42.4%) | 68 (50.7%) | 84 (48.8%) | 55 (41.0%) | 14 (8.1%) | 10 (7.5%) | 1 (0.6%) | 1 (0.7%) | 0,515 |
| **Chemical incident** | 67 (39.0%) | 73 (54.5%) | 83 (48.3%) | 52 (38.8%) | 19 (11.0%) | 9 (6.7%) | 3 (1.7%) | - | 0,022* |
| **Biological incident** | 64 (37.2%) | 64 (47.8%) | 79 (45.9%) | 58 (43.3%) | 23 (13.4%) | 9 (6.7%) | 6 (3.5%) | 3 (2.2%) | 0,124 |
| **Nuclear incident** | 56 (32.6%) | 54 (40.3%) | 82 (47.7%) | 59 (44.0%) | 27 (15.7%) | 14 (10.4%) | 7 (4.1%) | 7 (5.2%) | 0,355 |
| **Dirty bomb** | 57 (33.1%) | 59 (44.0%) | 82 (47.7%) | 52 (38.8%) | 28 (16.3%) | 18 (13.4%) | 5 (2.9%) | 5 (3.7%) | 0,227 |

*p<0,05 statistically significant.

**ONLINE SUPPLEMENTAL 6: PARTICIPANTS’ SELF-RATINGS ON THEIR PERCEIVED KNOWLEDGE, RISK AND DANGER OF DISASTERS**

| **Disaster scenario**  **(Median + IQR)** | **Knowledge** | **Risk** | **Danger** |
| --- | --- | --- | --- |
| **Natural disaster** | 5  (5-7) | 5  (3-5) | 5  (5-7) |
| **Bombing** | 5  (3-6) | 4  (2-5) | 7  (5-8,25) |
| **Influenza epidemic** | 8  (7-8) | 8  (7-9) | 5  (5-7) |
| **SARS-CoV-2 pandemic** | 8  (8-9) | 7  (5-8) | 6  (5-7,25) |
| **Mass-shooting** | 5  (2,75-6) | 4  (2-5) | 8  (5-9) |
| **Ebola/HF** | 4  (3-5) | 3  (2-5) | 8  (6-9) |
| **Smallpox** | 5  (3-5) | 4  (2-5) | 5  (5-7) |
| **Chemical incident** | 5  (3-6) | 5  (4-7) | 7  (5-8) |
| **Biological incident** | 4  (2-5) | 4  (2-5) | 7  (5-8) |
| **Nuclear incident** | 4  (2-5) | 5  (2-6) | 8  (7-9) |
| **Dirty bomb** | 3  (2-5) | 3  (2-5) | 8  (7-9) |

Rated on a 10-point scale (10 = highest, 1 = lowest).
